# Oxygen Uptake Efficiency Slope (OUES) and Mortality in Chronic Heart Failure: Comparisons of Absolute and Percent-predicted Values across Different Equations

**DOI:** 10.64898/2026.11.13.26367326

**Authors:** Gabriela Leopoldino Costa, Giovanni Donelli Costa, Fernando Colares Barros, Willian RobertoMenegazzo, Luiza Ferreira Van der Sand, Marina Petersen Saadi, Juliana Goulart Prata Oliveira Milani, Mauricio Milani, Anderson Donelli da Silveira

## Abstract

**Aim:** The oxygen uptake efficiency slope (OUES) is a prognostic indicator obtained from cardiopulmonary exercise testing (CPX) in chronic heart failure (CHF) patients. Although various equations exist for calculating predicted OUES, it remains unclear whether absolute or percent-predicted values offer a more accurate prognostic assessment. To investigate and compare mortality prediction in CHF patients using the OUES absolute values or the percent-predicted approach, incorporating the equations of Hollenberg, Buys, and Milani.

**Methods:** Retrospective cohort study involving CHF patients evaluated at a tertiary university hospital between July 2013 and July 2023. All patients underwent maximal CPX and were followed in a specialized outpatient clinic. The primary outcome was all-cause mortality. To compare the prognostic accuracy between different approaches, we performed receiver operating characteristic (ROC) curve analysis and Cox proportional hazards regression.

**Results:** A total of 1.180 patients were included, 57% male, with a mean age of 55 ± 12 years. The median follow-up was 5.0 years (IQR 3.0 to 7.2 years), resulting in 147 deaths (12.5%). The area under de curve (AUC) was 0.71 to 0.72 for males, and 0.60 to 0.63 for females, without statistical difference in the AUC between absolute and percent-predicted OUES. Different cut-offs were identified for each sex for predicting mortality and all OUES metrics remained independent predictors in multivariate models using sex and age.

**Conclusions:** OUES, in both absolute and percent-predicted forms, demonstrated similar accuracy in predicting mortality in CHF, with sex-specific cut-offs influencing clinical assessment.

**Condensed Abstract:** *Aim:* Compare absolute and percent-predicted OUES (Hollenberg, Buys, Milani) for mortality prediction in chronic heart failure.

*Methods:* Retrospective cohort of 1,180 patients undergoing maximal CPX.

*Results:* Both approaches showed similar prognostic accuracy, with sex-specific cut-offs.

*Conclusions:* Absolute and predicted OUES independently predicted mortality, supporting either approach for clinical risk stratification.

**Main Perspective:** Cardiopulmonary exercise testing plays a central role in risk stratification for patients with heart failure, and the oxygen uptake efficiency slope (OUES) is a robust prognostic marker even when maximal effort is not achieved. In this study of more than 1,100 patients, absolute and age- and sex-adjusted OUES showed comparable accuracy for predicting mortality. We also identified sex-specific OUES thresholds, supporting a more individualized approach to prognostic assessment and clinical decision-making in heart failure.

## Introduction

Heart Failure (HF) represents a major global public health challenge, being associated with significant functional impairment, reduced quality of life, and high healthcare resource utilization. It is estimated that more than 64 million people are currently living with this condition worldwide, reflecting its high prevalence and substantial socioeconomic impact (1–4). This leads to functional limitation, reduced exercise capacity, and poorer quality of life and survival, frequently manifested as poor performance in exercise testing (5–7). Disease progression accounts for most hospitalizations due to acute decompensation, with in-hospital mortality rates of 4–6% and one-year readmission and mortality rates ranging from 10–30% (8–10).

Cardiopulmonary exercise testing (CPX) is the gold standard for prognostic evaluation in cardiac and pulmonary diseases, allowing an integrated analysis of cardiovascular, ventilatory and metabolic responses to exertion. Parameters such as peak oxygen uptake (VO₂ peak), oxygen uptake efficiency slope (OUES), and the ventilatory equivalent for carbon dioxide (VE/VCO₂ *slope*) have gained increasing prominence (11,12). Among these, OUES stands out as a reliable marker of ventilatory efficiency and an important prognostic predictor in HF patients, especially due to its stability between maximal and submaximal exercise intensities (2,4,13). It represents an objective estimate of cardiorespiratory functional reserve, influenced by factors such as age, sex, and body composition, which justifies expressing it as a percentage of the predicted value for greater clinical precision (2,4).

OUES is also recognized as an independent prognostic predictor, particularly when combined with other indicators such as VO₂ peak, VE/VCO₂ *slope*, and anaerobic threshold (AT). Values below 1470 mL/min indicate worse prognosis in HF patients (12,14). Specific equations have been developed to predict OUES, notably those by Hollenberg (2), Buys (3), and Milani (4), the latter based on a South American sample to improve regional accuracy. However, a robust comparative analysis of the prognostic performance of absolute OUES values versus percent-predicted values in mortality prediction among HF patients is still lacking (5,6).

Thus, our objective was to investigate and compare mortality prediction in HF using absolute OUES values and the percent-predicted approach, incorporating the equations of Hollenberg (2), Buys (3), and Milani (4).

## METHODS

### Study design

We conducted a retrospective cohort study involving patients with chronic heart failure (CHF) undergoing CPX at a tertiary hospital in southern Brazil between July 2013 and July 2023. All examinations were performed during outpatient follow-up or in hospitalized patients with compensated HF before hospital discharge. For the current analysis, only the first CPX of each patient was used. Patients were instructed to maintain their current treatment. This study complies with the Declaration of Helsinki (6), with the research protocol having been approved by the local ethics committee, and informed consent obtained from all participants.

### Patients

We included individuals over 18 years of age with known HF. Subjects were clinically stable prior to CPX and were on optimized drug therapy. Exclusion criteria for those undergoing CPX were inability to perform the test due to motor or cognitive problems, orthopedic limitations, decompensated HF, acute infectious disease, and significant fluid and electrolyte disturbance. Patients were followed up periodically through outpatient visits at our institution. Clinical follow-up was assessed prospectively through electronic medical records and telephone calls. We considered patients lost to follow-up if they remained for at least 12 months without outpatient or emergency care at this hospital and who were not located by telephone contact.

### Cardiopulmonary exercise test

The exam was performed in the same room, in a monitored environment, with average temperatures of 18 to 22 °C. All tests were performed on a treadmill (GE T-2100, General Electric, USA) with breath-by-breath gas analysis (Metalyzer 3B, Cortex, Leipzig, Germany or Quark CPX, COSMED, Rome, Italy). Symptom-limited maximal exercise testing with an individualized ramp protocol was used to yield fatigue-limited exercise duration of 8 to 12 min. Maximal effort was considered when the respiratory exchange ratio was equal to or above 1.05.

Criteria for terminating the test were: patient desire, limiting angina, ST-segment elevation in an area without Q wave, supraventricular tachyarrhythmias with hemodynamic instability, sustained ventricular arrhythmias, pre-syncope or signs of low cardiac output. Methodological aspects of CPX follow previously validated recommendations, and all exams were executed by experienced and trained cardiologists using standardized institutional protocols.

Peak VO_2_ was determined by the highest measurement of a 20 s rolling average of breath-by-breath values. Percent-predicted VO_2_ was calculated using the Wasserman and Hansen algorithm. VE/VCO_2_ slope was determined by a linear regression model, with VE as the dependent variable (y-axis) and VCO_2_ as the independent variable (x-axis), using data from the entire duration of the test. OUES was defined as the gradient of the linear relationship of log10 minute ventilation (VE) to VO_2_. The OUES was measured throughout the CPX and calculated as described by Baba et. al using all exercise data (12).

The equations used to estimate OUES predicted values included: Hollenberg (2), who proposed a predictive equation for OUES based on a population sample from the United States, comprising individuals aged between 53 and 89 years; Buys (3), who presented absolute OUES values stratified by sex and age from a sample of healthy individuals in Europe (Belgium and the Netherlands), within the age range of 20 to 60 years; and Milani (4), who conducted a more comprehensive study with a large sample of healthy adults from South America, covering a broader age range (20 to 80 years). The present study not only compared the absolute OUES values by sex but also examined the percent-predicted values, providing a more comprehensive and adjusted perspective on OUES variability in relation to population factors. The formulas corresponding to the analyzed equations are presented in the supplementary appendix.

### Endpoints

The primary outcome was all-cause mortality. We performed receiver operating characteristic (ROC) curve analysis to compare the accuracy of the equations and absolute values and to determine the best cutoff points, according to optimal balance between sensitivity and specificity (Youden Index) (15).

### Statistical analysis

We used Shapiro Wilk test and histograms to assess normality of the variables. Continuous variables were compared by independent samples T-test and categorical variables by chi-square test. The area under the curve (AUC) of the different estimates was compared using the De Long method. Cox proportional hazards regression was employed to assess the univariate and multivariate effects of the various OUES metrics.

Statistical significance was defined as P < 0.05. All analyses were performed using R (R Foundation for Statistical Computing) or SPSS V.30 (IBM, Armonk,NY, USA).

## RESULTS

A total of 1.180 patients (679 males and 501 females) were analyzed, including 1.033 survivors and 147 non survivors. The mean age was slightly higher among patients who died (57.0 ± 12.1 years) compared to survivors (55.1 ± 11.9 years), with no statistically significant difference (p = 0.072). Male sex was more prevalent among survivors (p = 0.050).

Among comorbidities, significant differences were observed in the prevalence of diabetes mellitus (42.9% vs. 29.5%; p=0.002), atrial fibrillation (26.5% vs. 19.2%; p=0.047), chronic obstructive pulmonary disease – COPD (10.2% vs. 5.0%; p=0.020), and particularly chronic kidney disease (32.0% vs. 8.8%; p<0.001), all more frequent in the non-survivals.

Left ventricular ejection fraction (LVEF), was significantly lower among patients who died (31.4 ± 14.5% vs. 37.4 ± 15.4%; p<0.001), with a higher proportion of individuals with heart failure with reduced ejection fraction (HFrEF).

Regarding cardiopulmonary exercise testing parameters, patients who died presented lower VO₂ peak (15.4 ± 4.3 vs. 17.9 ± 4.9 mL/kg/min; p<0.001), a higher VE/VCO₂ slope (46.2 ± 12.2 vs. 39.0 ± 10.3; p<0.001), and lower OUES values (1,151 ± 418 vs. 1,462 ± 554; p<0.001), all with statistical significance, reinforcing the prognostic value of these functional variables.

The use of guideline-directed medical therapies was high in both groups, with no significant differences regarding the use of beta-blockers, ACE inhibitors or ARBs, spironolactone, or SGLT2 inhibitors (Table 1).

**Table 1.** Clinical and Demographic Characteristics of the Sample.

| Variable | Survivors<br>(N=1033) | Non-survivors<br>(N = 147) | p-value |
|---|---|---|---|
| Age, years | 55.12 (11.9) | 57.03 (12.1) | 0.072 |
| Male sex, n (%) | 583 (56.4%) | 96 (65.3%) | 0.050 |
| Female sex, n (%) | 450 (43.6%) | 51 (34.7%) | 0.050 |
| **Comorbidities (%)** |  |  |  |
| Hypertension | 523 (49.4%) | 71 (51.7%) | 0.598 |
| Diabetes | 728 (29.5%) | 84 (42.9%) | 0.002 |
| Atrial fibrillation | 835 (19.2%) | 108 (26.5%) | 0.047 |
| Dyslipidemia | 863 (16.5%) | 116 (21.1%) | 0.161 |
| Active smoking | 119 (11.9%) | 14 (9.9%) |  |
| Chronic kidney disease | 942 (8.8%) | 100 (32%) | <.001 |
| COPD | 981 (5.0%) | 132 (10.2%) | 0.020 |
| **LVEF (%)** | 37.37 (15.39) | 31.40 (14.50) | <.001 |
| HFrEF | 581 (65.1%) | 89 (80.9%) |  |
| HFmrEF | 95 (10.7%) | 7 (6.4%) |  |
| HFpEF | 144 (16.1%) | 11 (10%) |  |
| HFimpEF | 61 (6.8%) | 3 (2.7%) |  |
| **Implantable devices (%)** |  |  |  |
| Pacemaker | 22 (7.5%) | 2 (3.6%) |  |
| Implantable cardioverter defibrillator (ICD) | 114 (38.6%) | 19 (34.5%) |  |
| CRT with defibrillator (CRT-D) | 80 (27.1%) | 13 (23.6%) |  |
| **Pharmacological therapy (%)** |  |  |  |
| Beta-blocker | 89 (91.4%) | 10 (93.2%) | 0.528 |
| ACEI or ARB | 416 (59.7%) | 61 (58.5%) | 0.788 |
| Spironolactone | 399 (61.4%) | 69 (53.1%) | 0.059 |
| ARNI | 944 (8.6%) | 138 (6.1%) | 0.423 |
| SGLT2 inhibitor | 999 (3.3%) | 146 (0.7%) | 0.114 |
| **CPET variables** |  |  |  |
| Peak VO<sub>2</sub> (mL/kg/min) | 17.91 (4.92) | 15.44 (4.29) | <.001 |
| VE/VCO<sub>2</sub> slope | 38.99 (10.28) | 46.16 (12.23) | <.001 |
| OUES | 1462.18 (553.84) | 1151.30 (418.60) | <.001 |
| HR peak | 132 (26.19) | 121 (23.56) | <.001 |
| HR, % predicted | 78.66 (14.16) | 74.20 (14.04) | 0.486 |

HFrEF: heart failure with reduced ejection fraction; HFmrEF: heart failure with mildly reduced ejection fraction; HFpEF: heart failure with preserved ejection fraction; HFimpEF: heart failure with improved ejection fraction; COPD: chronic obstructive pulmonary disease; LVEF: left ventricular ejection fraction; CRT: cardiac resynchronization therapy; ICD: implantable cardioverter-defibrillator; CRT-D: cardiac resynchronization therapy defibrillator; ACEI: angiotensin-converting enzyme inhibitor; ARB: angiotensin receptor blocker;ARNI: sacubitril-valsartan;SGLT2: sodium/glucose cotransporter 2;HR: Heart hate.

ROC curve analysis demonstrated similar accuracy for the absolute OUES values and all three equations (Figure 1 and Table 2). For males, the AUC for the absolute OUES was 0.724, for Hollenberg was 0.713, for Buys was 0.721 and for Milani was 0.713, and there were no statistical differences among these AUC values. Females exhibited lower AUC values, but these differences were also not significant (Absolute: 0.633; Hollenberg: 0.613, Buys: 0.610 and Milani: 0.601). The absolute and percent-predicted OUES cut-points are shown in Table 2. Notably, for absolute OUES, distinct values were observed between sexes (1470 for males and 1150 for females), indicating different cut-offs for survival prediction (Figure 2).

**Figure 1:**
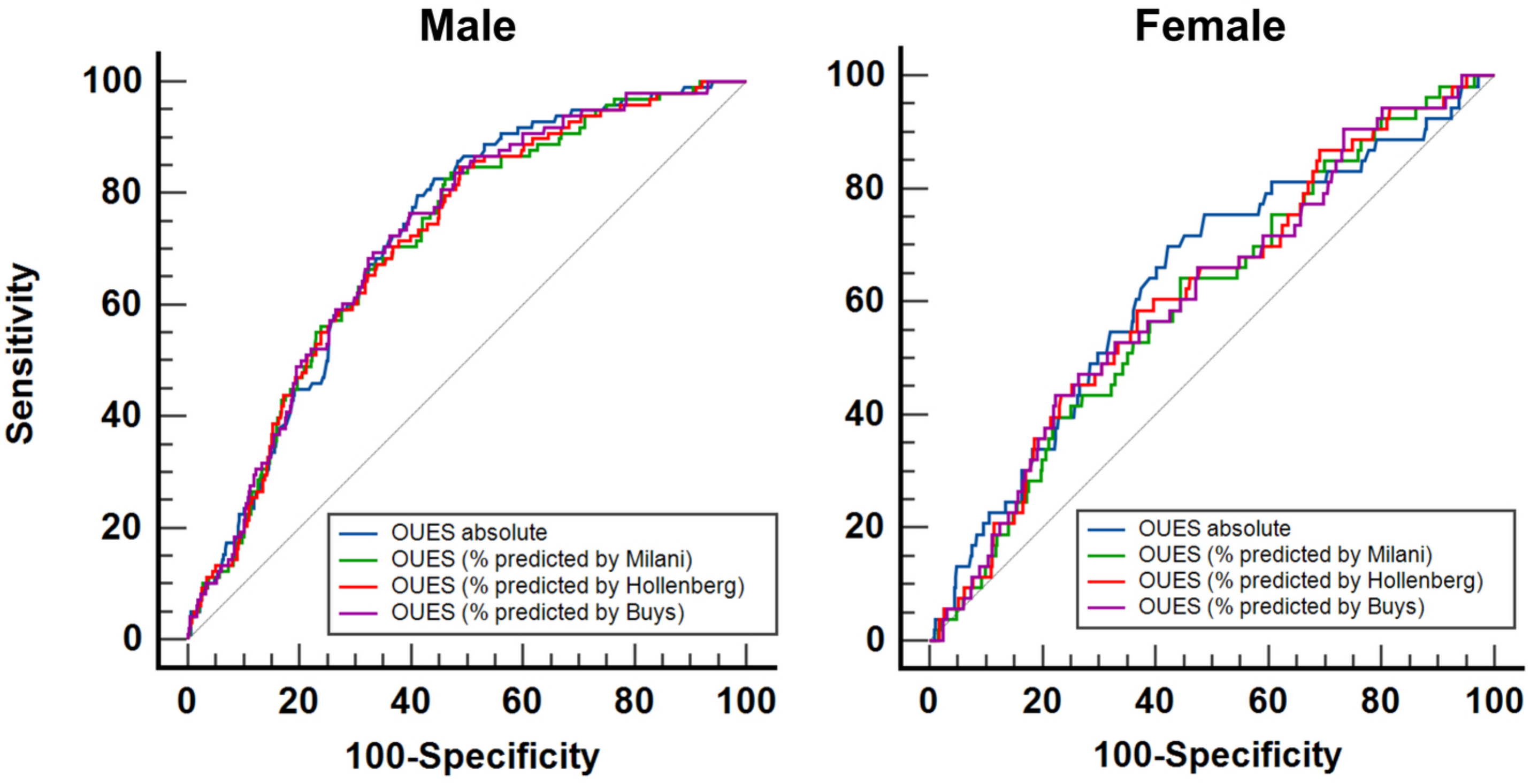
ROC curves for different measures of oxygen uptake efficiency slope (OUES) stratified by sex.

**Figure 2:**
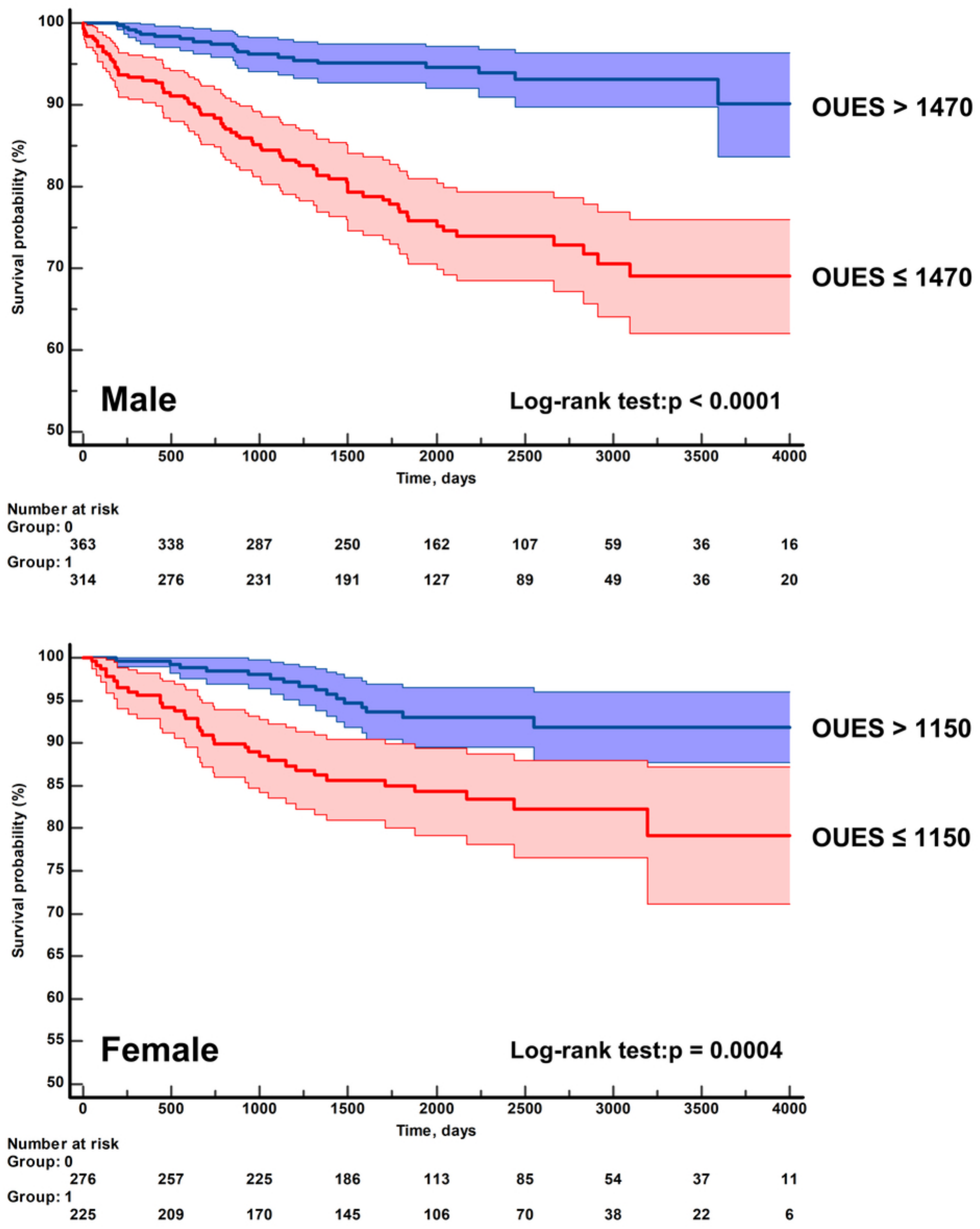
Kaplan-Meier curves for absolute oxygen uptake efficiency slope (OUES) stratified by sex.

**Table 2:** Absolute and percent-predicted values of OUES and its cut-points and accuracy across different sexes.

| Sex | Criteria | AUC and 95% CI | Cut-off (Youden) | Sensitivity | Specificity |
|---|---|---|---|---|---|
| Male | OUES absolute | 0.724 (0.688, 0.757) | ≤ 1470 | 79.6% | 58.8% |
|  | OUES Milani | 0.713 (0.677, 0.746) | ≤ 55.2% | 82.7% | 53.9% |
|  | OUES Holemberg | 0.713 (0.678, 0.747) | ≤ 62.6% | 84.7% | 51.3% |
|  | OUES Buys | 0.721 (0.686, 0.755) | ≤ 50.0% | 76.5% | 60.2% |
| Female | OUES absolute | 0.633 (0.589, 0.675) | ≤ 1150 | 69.8% | 57.8% |
|  | OUES Milani | 0.601 (0.556, 0.644) | ≤ 65.2% | 64.2% | 55.6% |
|  | OUES Holemberg | 0.613 (0.569, 0.656) | ≤ 64.1% | 58.5% | 63.1% |
|  | OUES Buys | 0.610 (0.566, 0.653) | ≤ 47.5% | 43.4% | 77.6% |

Statistics: Pairwise comparison of ROC curves by DeLong method did not reveal significant differences in both sexes.
AUC, area under the curve; CI, confidence interval; OUES, oxygen uptake efficiency slope.

In the survival analysis, absolute OUES had an unadjusted hazard ratio (HR) of 0.999, while percent-predicted OUES using Hollenberg had an HR of 0.972, Buys an HR of 0.967, and Milani an HR of 0.972 for mortality (P < 0.001 for all comparisons). Additionally, all OUES metrics remained significant, independent predictors after adjusting for age and sex in the multivariable models (P < 0.001) (Table 3).

**Table 3:**
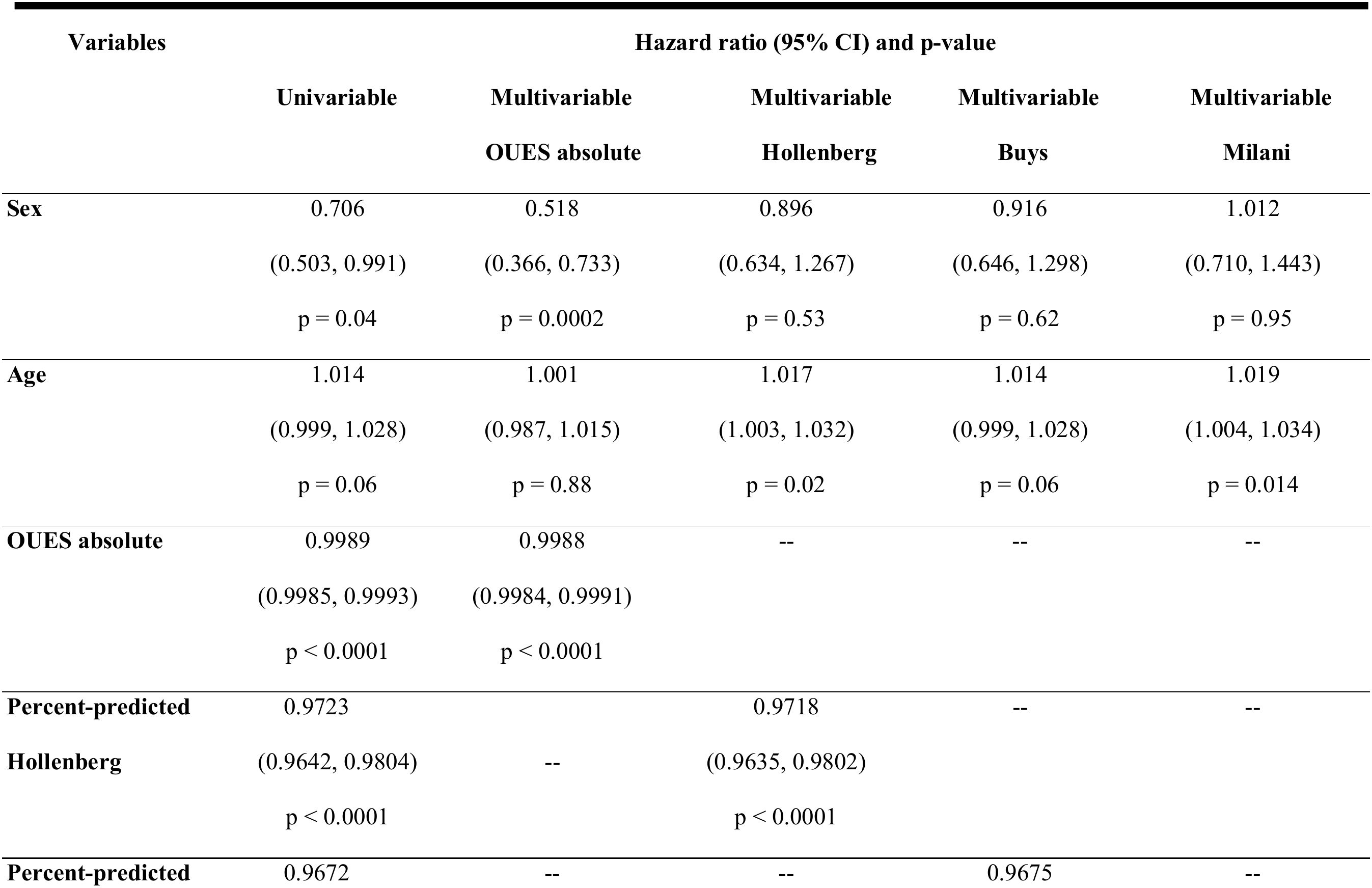
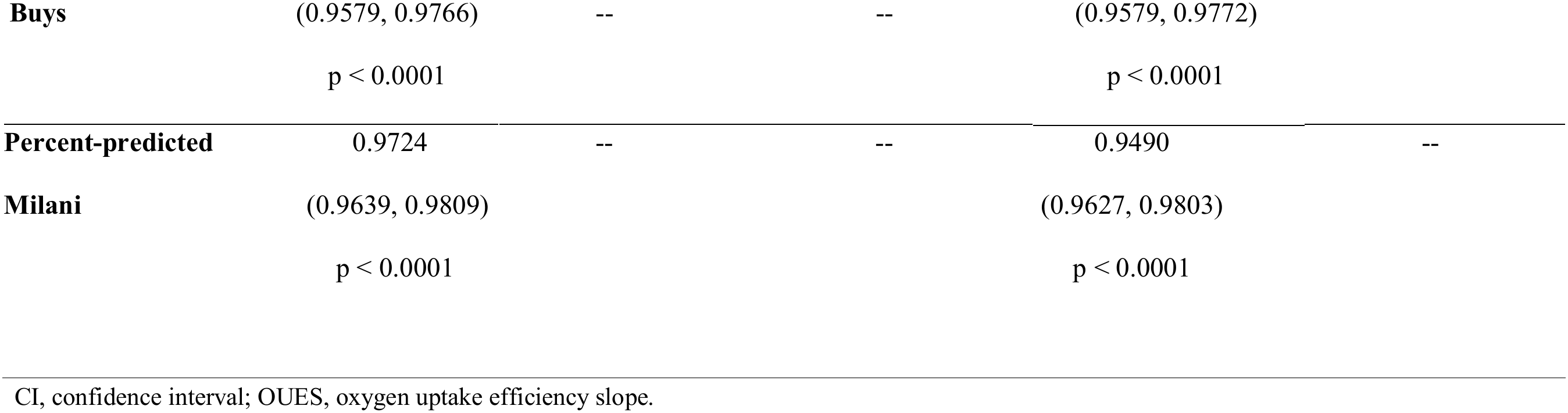
Cox regression models for Absolute and percent-predicted values for Oxygen Uptake Efficiency Slope (OUES)

| Variables | Hazard ratio (95% CI) and p-value |  |  |  |  |
|---|---|---|---|---|---|
|  | **Univariable** | **Multivariable<br>OUES absolute** | **Multivariable<br>Hollenberg** | **Multivariable<br>Buys** | **Multivariable<br>Milani** |
| **Sex** | 0.706<br>(0.503, 0.991)<br>p = 0.04 | 0.518<br>(0.366, 0.733)<br>p = 0.0002 | 0.896<br>(0.634, 1.267)<br>p = 0.53 | 0.916<br>(0.646, 1.298)<br>p = 0.62 | 1.012<br>(0.710, 1.443)<br>p = 0.95 |
| **Age** | 1.014<br>(0.999, 1.028)<br>p = 0.06 | 1.001<br>(0.987, 1.015)<br>p = 0.88 | 1.017<br>(1.003, 1.032)<br>p = 0.02 | 1.014<br>(0.999, 1.028)<br>p = 0.06 | 1.019<br>(1.004, 1.034)<br>p = 0.014 |
| **OUES absolute** | 0.9989<br>(0.9985, 0.9993)<br>p < 0.0001 | 0.9988<br>(0.9984, 0.9991)<br>p < 0.0001 | -- | -- | -- |
| **Percent-predicted<br>Hollenberg** | 0.9723<br>(0.9642, 0.9804)<br>p < 0.0001 | -- | 0.9718<br>(0.9635, 0.9802)<br>p < 0.0001 | -- | -- |
| **Percent-predicted<br>Buys** | 0.9672<br>(0.9579, 0.9766)<br>p < 0.0001 | --<br>-- | --<br>-- | 0.9675<br>(0.9579, 0.9772)<br>p < 0.0001 | -- |
| **Percent-predicted<br>Milani** | 0.9724<br>(0.9639, 0.9809)<br>p < 0.0001 | -- | -- | 0.9490<br>(0.9627, 0.9803)<br>p < 0.0001 | -- |

CI, confidence interval; OUES, oxygen uptake efficiency slope.

## DISCUSSION

Our findings indicate that the OUES, assessed by both absolute and percent-predicted values, demonstrates similar accuracy in predicting mortality in patients with CHF (16–17). The OUES stands out as a submaximal parameter of CPX, which evaluates the efficiency of oxygen uptake and utilization during progressive exercise, integrating ventilatory, cardiovascular and metabolic aspects that reflect the patient’s overall functional status. The sustained significance of the OUES after adjustment for age and sex reinforces its prognostic potential independent of these demographic factors, which is clinically relevant considering the heterogeneity of the CHF population. Furthermore, the robustness of the observed hazard ratios confirms that the OUES can be safely used in patients with exercise limitation, a common characteristic in this population.

It is important to highlight that a significant difference in the predictive ability of the OUES between men and women was observed, leading to the establishment of sex-specific cutoff points. This finding underscores the physiological and metabolic sexual differences that influence ventilatory response to exercise, indicating the need for sex-specific approaches in interpreting CPX data. Using a single cutoff point for both sexes may lead to under- or overestimation of risk, which directly impacts clinical management and prognostic stratification (17,18). Comparatively to other cohorts our study showed a higher percentage of female sex, which strengthens our sex-related findings.

Lewis et al. analyzed OUES and its prognostic role in CHF, being the first study to establish a cutoff value of 1.470 as the only significant independent prognostic variable, with 82% sensitivity and specificity, reinforcing its relevance in predicting clinical outcomes (13). Later, Myers et al. proposed a cutoff value of 1.400 for risk stratification in CHF patients.

However, our results highlight significant sex-based differences, suggesting that the OUES cutoff value for women may be considerably lower than that observed in men. These findings underscore the importance of personalized approaches in interpreting CPX data, as applying a single standardized cutoff across the entire population may compromise the diagnostic and prognostic accuracy of this parameter (19, 20).

Ross et al. used an equation Hollemberg in a study with 721 CHF patients, 82% of whom were men, but did not explore sex differences. By comparing the prognostic characteristics of absolute and percent-predicted OUES, they demonstrated its important prognostic value in CHF (22). Our analysis also revealed higher AUC values in men, in absolute and percent-predicted values, indicating a greater prognostic capacity of OUES in this group. This finding might be attributable to the observed higher mortality rate in males than females in our cohort (14.4 versus 10.5%).

In a retrospective cohort study including 128 patients with end-stage heart failure who underwent CPX, 43 experienced cardiac events (5 deaths and 38 urgent heart transplants) over 2 years. OUES ≥ 1.6 and diuretic use were predictors of survival, while peak VO₂ and VE/VCO₂ slope were not. Patients with higher OUES had better survival rates (p < 0.001). The authors concluded that OUES may aid in prognostic assessment of end-stage heart failure patients and be useful for evaluating those unable to perform maximal exercise testing (23).

The indiscriminate use of a universal cutoff point in heterogeneous populations may lead to misinterpretations - underestimating risk in individuals with inherently lower functional capacity, such as women, or overestimating it in other subgroups. A study involving 2,220 apparently healthy adults (48% women), with a mean age of 44.7 ± 12.9 years, who underwent CPX, analyzed OUES calculated across the entire test, as well as during the initial 50% (OUES 50) and 75% (OUES 75) of test duration, and normalized for body surface area. OUES, particularly in its normalized and OUES 75% forms, was associated with lower mortality in men, but not in women after adjusting for risk factors (20).

Therefore, in terms of risk stratification, the prognostic value of absolute OUES has been widely documented in apparently healthy men (3,4) and across various cardiovascular conditions (1). These findings highlight the potential of both absolute and percent-predicted OUES as valuable tools for mortality prediction in HF (11,19).

The equations analyzed by Hollenberg (2), Buys (3), and Milani (4) indicate that there is no significant difference when using the absolute value of OUES. This suggests that the results obtained are consistent, regardless of the methodology adopted for calculating OUES as a prognostic value and physical performance indicator in patients with CHF. Therefore, our study reinforces the need to reevaluate OUEScutoffs, proposing the adoption of sex-specific values. This personalized approach could improve the accuracy of risk stratification and provide a more robust basis for the clinical management of CHF patients, benefiting both men and women more equitably.

### Limitations

This study has several important limitations to consider when interpreting the results. First, as an observational, single-center study, its findings have limited generalizability and carry a higher risk of uncontrolled confounding due to patient heterogeneity. Second, referral for CPX was not standardized and relied on clinical judgment, which may have introduced indication bias. Additionally, losses to follow-up could have led to underestimation of adverse events, such as out-of-hospital deaths or hospitalizations elsewhere, affecting outcome accuracy.

## CONCLUSION

The OUES, assessed by absolute and percent-predicted values adjusted for age and sex, showed similar accuracy in predicting mortality in heart failure. This equivalence reinforces its clinical utility as a prognostic marker, even without maximal effort during CPX. Sex-specific cut-offs enhance precision in evaluating ventilatory efficiency and risk stratification, considering physiological differences between men and women. These findings establish OUES as a practical and sensitive tool for functional and prognostic assessment in heart failure.

## Supporting information

Manuscript

## Data Availability

The data that support the findings of this study are available from the corresponding author upon reasonable request. The data are not publicly available due to privacy and ethical restrictions.

## Acknowledgments

The authors thank the service teams for their technical and scientific support during data collection and analysis. We also express our gratitude to the patients who voluntarily participated in this study, making it possible to advance knowledge in the field.

## References

1. Savarese G, Becher PM, Lund LH, Seferovic P, Rosano GMC, Coats AJS. Global burden of heart failure: a comprehensive and updated review of epidemiology. Cardiovascular Research, V.118, 17, Dec.2022, Pag.3272–3287.

2. Hollenberg M, Tager IB. Oxygen uptake efficiency slope: an index of exercise performance and cardiopulmonary reserve requiring only submaximal exercise. JAm Coll Cardiol. 2000;36(1):194–201.

3. Buys R, Coeckelberghs E, Vanhees L, Vanhees L, Cornelissen VC. The oxygen uptake efficiency slope in 1411 Caucasian healthy men and women aged 20-60 years: reference values. Eur J Prev Cardiol. 2015;22(3):356–63.

4. Milani J, Milani M, Cipriano GFB, Castro I, Hansen D, Junior GC. Oxygen Uptake Efficiency Slope in South American Healthy Adults: comprehensive reference values and international comparisons. J Cardiopulm Rehabil Prev. 2023;43(4):290–300.

5. Heidenreich PA, Albert NM, Allen LA, Bluemke DA, Butler J, Fonarow GC, et al. Forecasting the impact of heart failure in the united states a policy statement from the american heart association. Circ Hear Fail. 2013;6(3):606–19.

6. WORLD MEDICAL ASSOCIATION. WMA Declaration of Helsinki: ethical principles for medical research involving human subjects. Adopted by the 18th WMA General Assembly, Helsinki, Finland, June 1964, and amended by the 75th WMA General Assembly, Helsinki, Finland, October 2024. Ferney-Voltaire: World Medical Association,2024.Disponível em:https://www.wma.net/policies-post/wma-declaration-of-helsinki-ethical-principles-for-medical-research-involving-human-subjects/. Acesso em: 15 jun. 2025.

7. Albuquerque DC, de Neto JD S, Bacal F, Rohde LEP, Bernardez-Pereira S, Berwanger O, et al. I registro Brasileiro de insuficiência Cardíaca – Aspectos clínicos, qualidade assistencial e desfechos hospitalares. Arq Bras Cardiol. 2015;104(6):433–42.

8. Solomon SD, Dobson J, Pocock S, Skali H, McMurray JJV, Granger CB, et al. Influence of nonfatal hospitalization for heart failure on subsequent mortality in patients with chronic heart failure. Circulation. 2007;116(13):1482–7.

9. Rohde LEP, Montera MW, Bocchi EA, Clausell NO, de Albuquerque DC, Rassi S, et al. Diretriz brasileira de insuficiência cardíaca crônica e aguda. Arq Bras Cardiol. 2018;111(3):436–539. Chin MH, Goldman L. Factors Contributing to the Hospitalization of Patients with Congestive Heart Failure. 1996;(February 1993):643–8.

10. Kaminsky LA, Arena R, Myers J, et al. Updated reference standards for cardiorespiratory fitness measured with cardiopulmonary exercise testing: data from the Fitness Registry and the Importance of Exercise National Database (FRIEND). Mayo Clin Proc. 2022;97(2):285–293.

11. Baba R, Nagashima M, Goto M, Nagano Y, Yokota M, Tauchi N, Nishibata K. Oxygen uptake efficiency slope: a new index of cardiorespiratory functional reserve derived from the relation between oxygen uptake and minute ventilation during incremental exercise. Journal of the American College of Cardiology. Volume 28, issue 6, 15 november 1996, pages 1567–1572.

12. Lewis CD, Roland W, Panagiota G, Mariantonietta C, Andrew J.S.C, Massimo F. et al. Enhanced prognostic value from cardiopulmonary exercise testing in chronic heart failure by non-linear analysis: oxygen uptake efficiency slope. European Heart Journal 2006 27, 684–690.

13. Wei-Jen C, Jen-Ting L, Su-Ying H, Po-Chen H, Chen-Liang C.Prognostic Value of Body Surface Area-Adjusted Oxygen Uptake Efficiency Slope in Heart Failure Patients. Acta Cardiol Sin. 2024 May;40(3):322–330.

14. Fluss, R.; Faraggi, D.; Reiser, B. Estimation of the Youden Index and its associated cutoff point.Biom J., v. 47, n. 4, p. 458–472, ago. 2005. DOI:10.1002/bimj.200410135.

15. Allen LA, Stevenson LW, Grady KL, Goldstein NE, Matlock DD, Arnold RM, et al. Decision making in advanced heart failure: A scientific statement from the american heart association. Circulation. 2012;125(15):1928–52.

16. Van Laethem C, Bartunek J, Goethals M, Nellens P, Andries E, Vanderheyden M. Oxygen uptake efficiency slope, a new submaximal parameter in evaluating exercise capacity in chronic heart failure patients. Am Heart J. 2005;149(1):175–180.

17. Kaminsky LA, Arena R, Myers J, et al. Updated reference standards for cardiorespiratory fitness measured with cardiopulmonary exercise testing: data from the Fitness Registry and the Importance of Exercise National Database (FRIEND). Mayo Clin Proc. 2022;97(2):285–293.

18. Myers J, Ross A, Frederick D, Daniel B, Joshua A, Leon H, Paul C, Marco G, Mary AP. A cardiopulmonary exercise testing score for predicting outcomes in patients with heart failure. Am Heart. 2008 Dec;156(6):1177–83.

19. Peterman JE, Novelli DS, Fleenor BS, Whaley MH, Kaminsky LA, Harber MP. Oxygen uptake efficiency slope as a predictor of mortality risk: the ball state adult fitness longitudinal lifestyle study (BALLST) published online ahead of print J Cardiopulm Rehabil Prev.2023 July 1;43(4):282–289.

20. Lima RM, Vainshelboim B, Ganatra R, Dalman R, Chan K, Myers J. Exercise training improves ventilatory efficiency in patients with a small abdominal aortic aneurysm: a randomized controlled study. J Cardiopulm Rehabil Prev. 2018;38(4):239–245.

21. Ross Arena, Jonathan Myers, Joshua Abella, Sherry Pinkstaff, Peter Brubaker, Dalane Kitzman, Mary Ann Peberdy, Daniel Bensimhon, Paul Chase, Med, Marco Guazzi Prognostic Significance of the Oxygen Uptake Efficiency Slope: Percent-Predicted versus Actual Value. Am J Cardiol. 2010 March 1; 105(5): 757–758.

22. Huang C, Chen JY, Chen HC, Huang CW, Lin LK. The Pre-Discharge Oxygen Uptake Efficiency Slope Predicts One-Year Cardiovascular Events in Acute Decompensated Heart Failure Patients. Life (Basel). 2022 Sep 19;12(9):1449.

