## Supplementary material for "Oxygen Uptake Efficiency Slope (OUES) and Mortality in Chronic Heart Failure: Comparisons of Absolute and Percent-predicted Values across Different Equations": Manuscript

**Supplementary appendix**

### Equations used for predicted Oxygen Uptake Efficiency Slope (OUES)

#### Hollenberg Equation:

Females:

$OUES=1,175-(15.8 \cdot age)+(851 \cdot BSA)$

Males:

$OUES=1,320-(26.7 \cdot age)+(1,394 \cdot BSA)$

#### Buys Equation:

Females:

$842-18.5*age + 1280* BSA$

Males:

$1093 - 18.5* age + 1479* BSA$

#### Milani Equation:

Females:

$OUES=-1738+(34.18 \times Age)-(0.4949 \times Age2)+(8.561 \times Weight)+(1650 \times Height);r2 =0.174$

Males:

$OUES = -1786+(30.95 \times Age)-(0.5389 \times Age 2 )+(14.12 \times Weight)+(2016 \times Height);r2 =0.239$

#### Reference:

1. Hollenberg M, Tager IB. Oxygen uptake efficiency slope: an index of exercise performance and cardiopulmonary reserve requiring only submaximal exercise. JAm Coll Cardiol. 2000;36(1):194-201.

2. Buys R, Coeckelberghs E, Vanhees L, Vanhees L, Cornelissen VC. The oxygen uptake efficiency slope in 1411 Caucasian healthy men and women aged 20-60 years: reference values. Eur J Prev Cardiol. 2015;22(3):356-63.

3. Milani J, Milani M, Cipriano GFB, Castro I, Hansen D, Junior GC. Oxygen Uptake Efficiency Slope in South American Healthy Adults: comprehensive reference values and international comparisons. J Cardiopulm Rehabil Prev. 2023;43(4):290-300.
